# Tumescent Local Anesthesia: A Systematic Review of Outcomes

**DOI:** 10.1101/2020.08.10.20170720

**Authors:** Yu Liu, Sanjana Lyengar, Chrysalyne D Schmults, Emily S Ruiz, Robert Besaw, Laura K Tom, Michelangelo Giovanni Vestita, Jason Kass, Abigail H Waldman

## Abstract

**IMPORTANCE:** Tumescent local anesthesia (TLA, whereby anesthesia is achieved by injection of a highly diluted solution of local anesthesia into skin and subcutaneous tissues) is a technique for delivering anesthesia for superficial surgical procedures. TLA obviates the need for general anesthesia or intravenous sedation in most cases. Pain control and TLA-related complications are key factors in determining the success of TLA.

**OBJECTIVE:** To conduct a systematic review of the English medical literature’s data regarding pain control and TLA-related complications in TLA surgical cases to determine its efficacy and safety

**EVIDENCE REVIEW:** The review was performed in accordance with the Preferred Reporting Items for Systematic Reviews and Meta-Analyses guidelines (PRISMA). Searches of both the MEDLINE and EMBASE databases were performed. Articles using 10-point quantitative scales were included in the pain analysis. Complications were tabulated from cohort studies, case series, and case reports. A total of 184 articles cotaining reports of 71,483 surgical procedures met inclusion criteria, including 43 with pain outcomes and 141 reporting complications.

**FINDINGS:** Liposuction procedures were associated with relatively low degree of both intraoperative pain (10-point visual analog scale 1.1 ± 2.1) and post-operative pain (0.53 ± 0.44) and the fewest complications (1.2%). The highest intra-operative and post-operative pain was reported in facial/cleft-lip surgery (3.7 and 3.99, respectively), while mastectomy was associated with highest post-operative complication risk (20.8%). There were 8 reported cases of death unlikely related to TLA: pulmonary embolus (4 cases), complications related to concurrent general anesthesia (2 cases), hemorrhage, and visceral perforation. There were 5 reported cases of death related to TLA (lidocaine/bupivacaine toxicity in 4 cases and one case of fluid overload) during its development when optimal dose and volume parameters were being established. There have been no TLA-associated deaths reported in the 33,429 cases published since 2003.

**CONCLUSIONS AND RELEVANCE:** This systematic review demonstrates TLA to be a safe and effective anesthetic approach. Its low-cost and rapid patient recovery warrant further studies of cost-reduction and patient satisfaction. Expanded education of TLA techniques in surgical and anesthesia training programs may be considered to broaden patient access to this anesthetic modality for cutaneous and subcutaneous surgical procedures.

**Key Points:** *Question:* Is TLA an effective and safe local anesthetic technique for pain management during surgical procedures?

*Findings:* In this review of 157 publications, TLA was a safe and effective anesthetic approach. The least pain and fewest complications were in liposuction procedures. The highest postoperative complication risk was with mastectomy. Though five TLA-related deaths were reported in early liposuction cases, there have been no deaths in the 33,429 TLA cases published since 2003.

*Meaning:* TLA is an effective and safe anesthetic technique which enables cutaneous and subcutaneous surgery to be performed in office-based settings with high safety and low cost.

## INTRODUCTION

Tumescent local anesthesia (TLA) is an anesthetic technique which uses the infiltration of a very dilute local anesthetic (usually lidocaine) into tissue to achieve an anesthetic effect while minimizing side effects due to the anesthetic components.^1-5^ The makeup of TLA has historically consisted of a diluted local anesthetic combined with epinephrine and sodium bicarbonate, for example, 500-1000 mg lidocaine and 1 mg epinephrine solution and 12.5 meq sodium bicarbonate solution with 1000 ml normal saline^2^ Since Jeffrey Klein first introduced this method to the field of dermatologic anesthetic surgery in the 1980s,^1,2^ TLA has gained widespread interest within the medical community.^4,5^ Its application has since expanded to include breast reconstruction, hand, and burn surgeries among others due to its many benefits.^4-12^ Potential advantages of utilizing TLA include: 1) low reported incidence of post-operative complications,^9^ 2) reduced blood loss due to epinephrine-induced vasoconstriction and hydrostatic compression from the tumescent effect,^2,13^ 3) low infection rates due to potential bacteriostatic effects of lidocaine and common use of outpatient surgical environments,^14,15^ 4) pain relief from the alkaline component of the TLA solution,^13,16-19^ 5) relatively long lasting anesthetic effects,^20-22^ and 6) lower costs when compared to general anesthesia.

Over the last three decades, the specific local anesthetic agents used have been expanded from classical lidocaine to other amino amide or amino ester anesthetics, including articaine, bupivacaine, etidocaine, mepivacaine and prilocaine amide class, as well as chloroprocaine, procaine, and tetracaine ester class anesthetics.^4,5^ Of note, lidocaine is currently still used in the vast majority of TLA applications.^4,5^ In current liposuction procedures, lidocaine (with epinephrine) in doses 35-55 mg/kg is routinely used with an excellent safety record.^2,5,23^ Klein et al, 2016 notes that the liposuction procedure itself may remove a portion of the TLA lidocaine during suction, before the drug can be absorbed into the systemic circulation.^24^ Therefore, the optimal dose of TLA lidocaine for procedures other than liposuction may be lower. Recent literature suggests that the maximum safe dosages of TLA lidocaine may be 28 mg/kg for procedures without liposuction,^5,24^ and 45 mg/kg for procedures with liposuction.^24^

A number of review articles have been written on TLA focusing on technique.^4,5,8-10^ However, none have systematically summarized the available data regarding pain control and postoperative complications - two key factors in determining the success of an anesthesia protocol. In the current work, the published English literature reporting pain outcomes and complications has been summarized and analyzed, including studies that directly compare TLA to general anesthesia (GA), in order to provide physicians with a comprehensive, up-to-date summary of the current uses, safety, and efficacy of TLA as compared to GA.

## METHODS

### Search strategy

A systematic review was performed in accordance with the Preferred Reporting Items for Systematic Reviews and Meta-Analyses guidelines [PRISMA citation] to evaluate all available data published in English on pain and procedure-related complications relating to TLA use in surgical procedures. Searches of both the MEDLINE and EMBASE databases were performed on June 1, 2017 and August 20, 2019. The following search terms were used for study extraction: “*xylocaine tumescent”, “lidocaine tumescent”, “xylocaine dilution”, “lidocaine dilution” “xylocaine dilutional”, “lidocaine dilutional”, “tumescent”, “tumescence anesthesia”, “tumescence local”*.

### Selection criteria

Only articles where procedures were completed using TLA were included. Post and peri-operative complications, regardless of whether they were attributed to TLA, were tabulated from cohort studies, case series, and case reports. Articles that reported pain data using a quantitative scale (VAS, visual analog scale) were included in the pain analysis.

### Study selection and data extraction

All articles yielded by the aforementioned search were screened independently for eligibility by two researchers. Reference lists from articles were also reviewed to identify additional articles that were missed in the initial screen. Studies containing primary data regarding pain or complications were included. The following post-operative complication parameters were extracted from each article: presence of hematoma, seroma, infection, severe scarring, nerve injury, skin and/or fat necrosis at the procedure site, wound dehiscence, and any other adverse post-operative sequelae. Primary intra-operative and post-operative pain data that recorded pain based on the visual analog scale (VAS) or an equivalent (0-10 point scoring system with 0 being no pain and 10 being the worst possible pain) were included in the pain analysis.

### Statistical analysis

Statistical analyses were conducted using SPSS statistical software. Complication outcomes were tabulated as the number of total incidents in each parameter, and the percentage of patients experiencing each parameter. Pain data were summarized as weighted means and standard deviations of the reported VAS scores for each procedure type. Pain calculations were weighted according to number of patients in each procedure category.

## RESULTS

The searches of MEDLINE and EMBASE yielded 3,336 articles (Figure 1). Duplicates and nonrelevant papers were removed, leaving 737 articles to be screened. Ultimately, 553 articles were excluded because they failed to meet selection criteria or did not contain the required information as indicated in Figure 1. Of the remaining 157 articles, 32 relating to pain outcomes and 125 relating to post-operative complication outcomes were included for analysis. The numbers of published works in liposuction, cutaneous surgery, plastic surgery, breast procedures, and hand surgery are reported in Figure 2.

**Figure 1.**
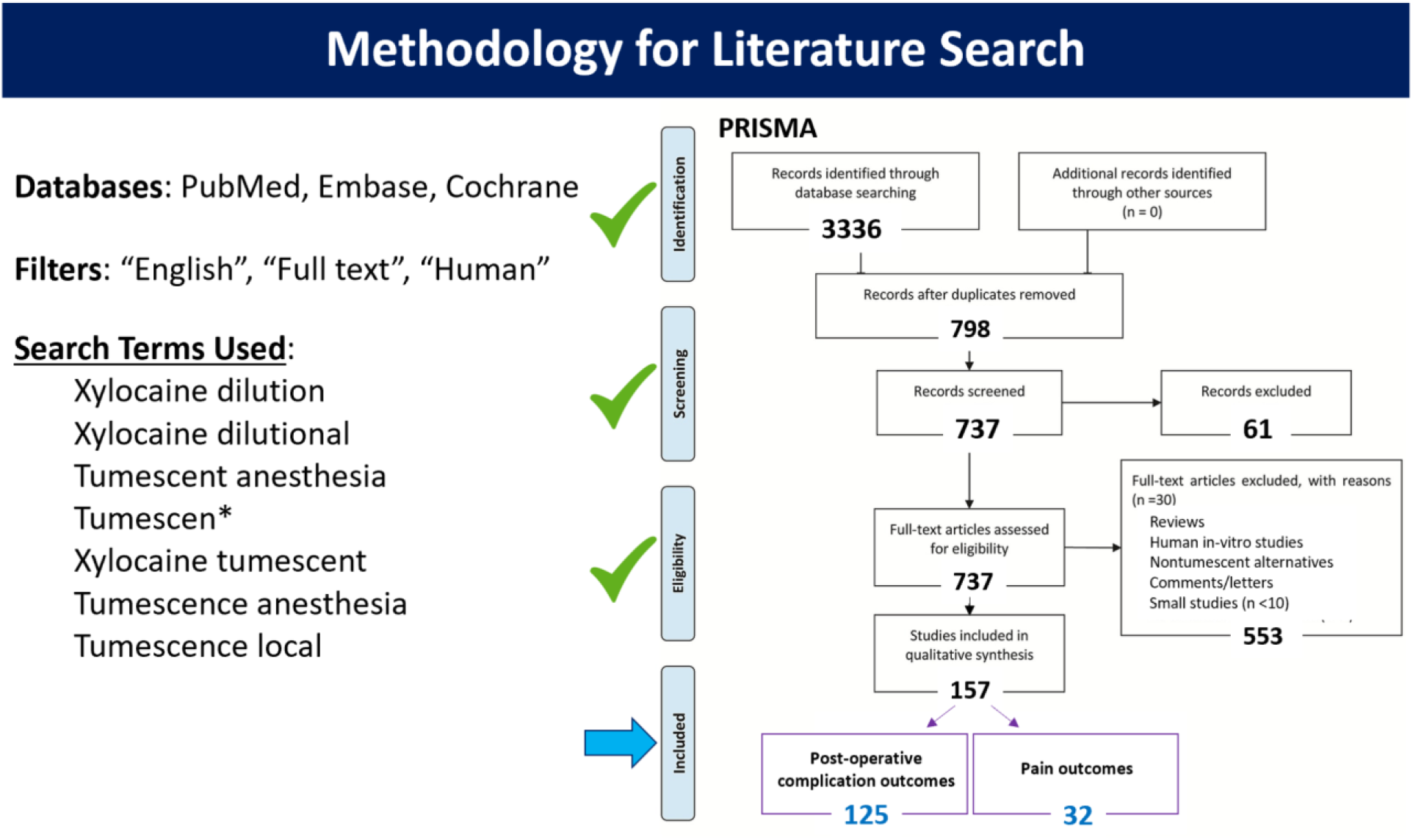
Flow chart summary of search process.

**Figure 2.**
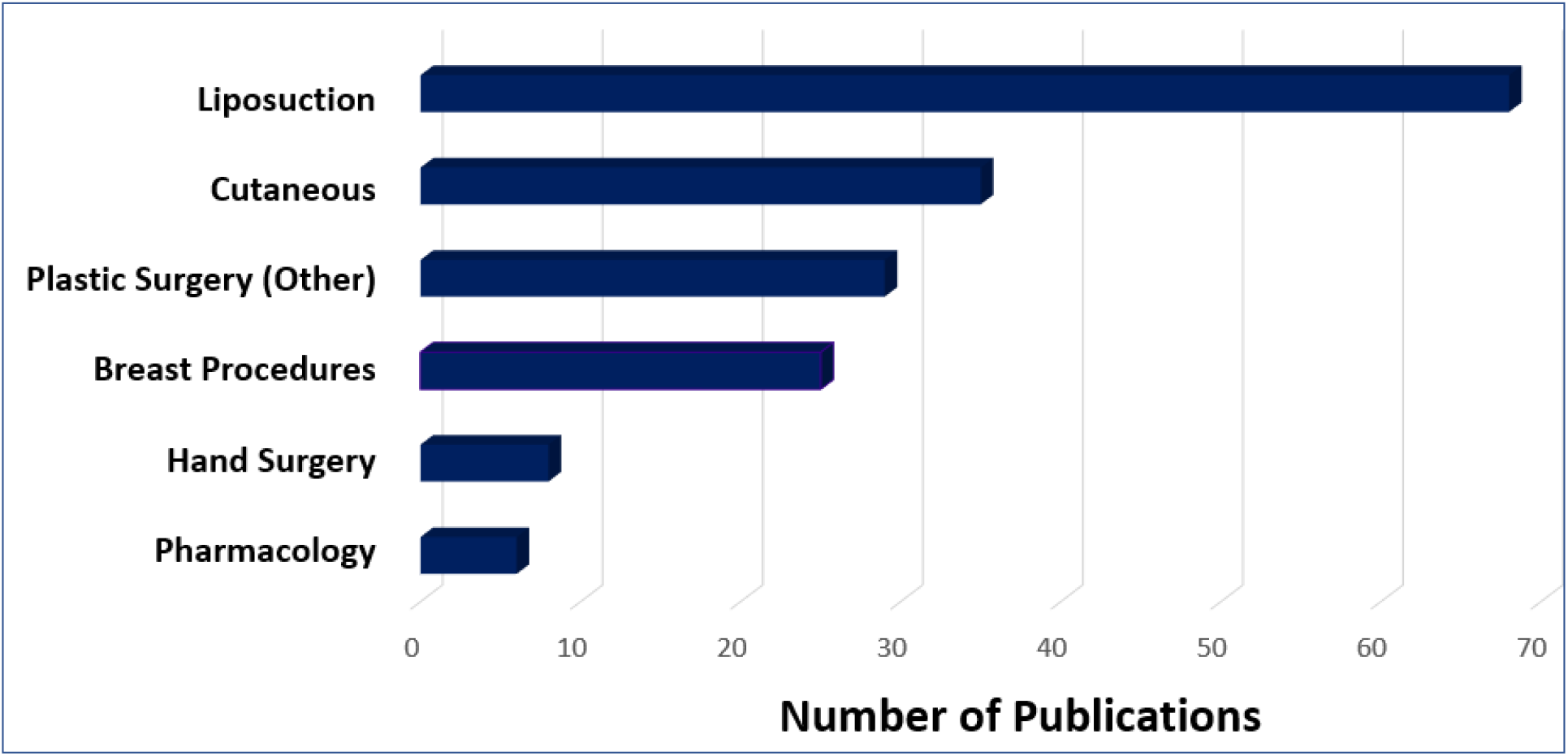
Number of TLA publications by procedure type.

### Intra-operative Pain Outcomes (Table 1)

To quantify the amount of pain during TLA procedures, intra-operative pain data from 15 published works was analyzed (see Table 1). All pain data were reported using the visual analog scale (VAS) system or a comparable quantitative pain scale. Pain scores were reported after injection of TLA at various time points during execution of vascular, plastic or hand surgical procedures (Table 1).

**Table 1.**
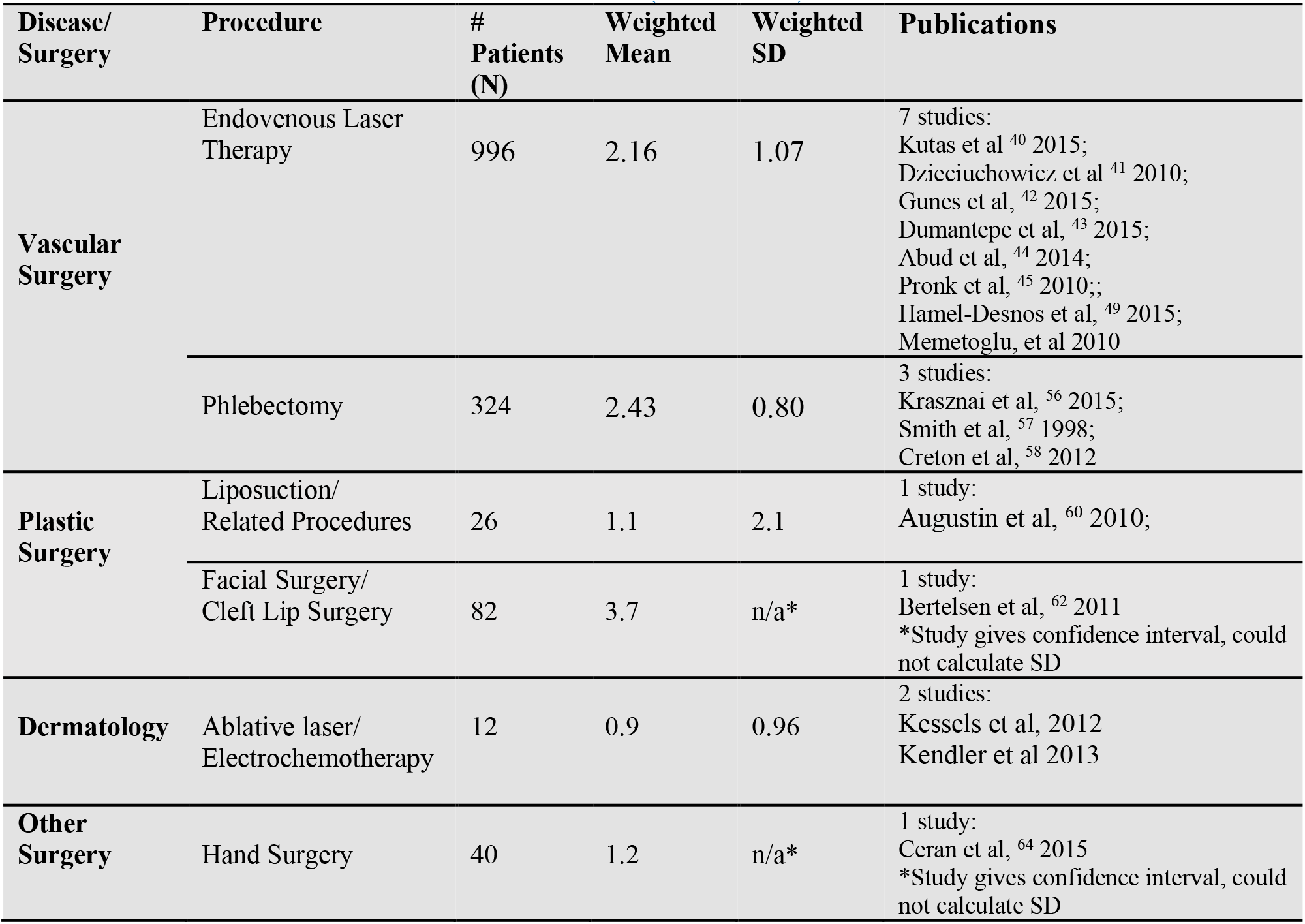
INTRA-OPERATIVE PAIN DATA (VAS, 0-10)

| **Disease/**<br>**Surgery** | **Procedure** | **#**<br>**Patients**<br>**(N)** | **Weighted**<br>**Mean** | **Weighted**<br>**SD** | **Publications** |
| --- | --- | --- | --- | --- | --- |
| **Vascular**<br>**Surgery** | Endovenous Laser<br>Therapy | 996 | 2.16 | 1.07 | 7 studies:<br>Kutas et al <sup>40</sup> 2015;<br>Dzieciuchowicz et al <sup>41</sup> 2010;<br>Gunes et al, <sup>42</sup> 2015;<br>Dumantepe et al, <sup>43</sup> 2015;<br>Abud et al, <sup>44</sup> 2014;<br>Pronk et al, <sup>45</sup> 2010;;<br>Hamel-Desnos et al, <sup>49</sup> 2015;<br>Memetoglu, et al 2010 |
|  | Phlebectomy | 324 | 2.43 | 0.80 | 3 studies:<br>Krasznai et al, <sup>56</sup> 2015;<br>Smith et al, <sup>57</sup> 1998;<br>Creton et al, <sup>58</sup> 2012 |
| **Plastic**<br>**Surgery** | Liposuction/<br>Related Procedures | 26 | 1.1 | 2.1 | 1 study:<br>Augustin et al, <sup>60</sup> 2010; |
|  | Facial Surgery/<br>Cleft Lip Surgery | 82 | 3.7 | n/a* | 1 study:<br>Bertelsen et al, <sup>62</sup> 2011<br>*Study gives confidence interval, could<br>not calculate SD |
| **Dermatology** | Ablative laser/<br>Electrochemotherapy | 12 | 0.9 | 0.96 | 2 studies:<br>Kessels et al, 2012<br>Kendler et al 2013 |
| **Other**<br>**Surgery** | Hand Surgery | 40 | 1.2 | n/a* | 1 study:<br>Ceran et al, <sup>64</sup> 2015<br>*Study gives confidence interval, could<br>not calculate SD |

Dermatologic procedures and liposuction procedures were associated with the lowest degree of intra-operative pain with a mean of 0.9 for dermatologic procedures (Standard deviation, SD 0.96), and 1.1 (SD 2.1) for liposuction procedures. Weighted mean pain scores during facial/cleft-lip surgery were 3.7 (original SD data not provided). Varicose vein treatments were associated with relatively low pain: 2.16 (SD 1.07) for endovenous laser treatment (EVLT/ELT) and 2.43 (SD 0.80) for phlebectomy.

### Post-operative Pain Outcomes (Table 2)

Data on post-operative pain (within the first post-operative day) was gathered from 28 published works (Table 2). The lowest post-operative pain scores were reported for liposuction procedures 0. 53 (SD 0.44), followed by radiofrequency ablation therapy was 0.65 (0.10) for treatment of varicose veins. The weighted mean VAS score for other vascular surgery were medium, with phlebectomy and endovenous laser therapy procedures 1.73 (SD 0.52) and 2.44 (SD 1.35), respectively. The weighted mean VAS score were 2.6 (SD not provided) for breast procedures, 2.77 (SD 0.43) for hand surgery, then facial/cleft-lip procedures 3.99 (SD 0.49).

**Table 2.** POST-OPERATIVE DAY ONE PAIN DATA (VAS, 0-10)

| **Disease/**<br>**Surgery** | **Procedure** | **# Patients**<br>**(N)** | **Weighted**<br>**Mean** | **Weighted**<br>**SD** | **Publications** |
| --- | --- | --- | --- | --- | --- |
| **Vascular**<br>**Surgery** | Endovenous<br>Laser Therapy | 1466 | 2.44 | 1.35 | 13 studies:<br>Gunes et al, <sup>42</sup> 2015;<br>Dumantepe et al, <sup>43</sup> 2015;<br>Abud et al, <sup>44</sup> 2014;<br>Pronk et al, <sup>45</sup> 2010;;<br>Jibiki et al, <sup>47</sup> 2016);;<br>Memetoglu et al, <sup>48</sup> 2010;<br>Hamel-Desnos et al, <sup>49</sup> 2015;<br>Pannier et al, <sup>50</sup> 2011;<br>Van Zandvoort et al, <sup>51</sup> 2016;<br>Wallace et al, <sup>52</sup> 2017;<br>Rasmussen et al, <sup>53</sup> 2007<br>Nandhra et al, 2018<br>Wahbi et al, 2017 |
|  | Radiofrequency<br>Ablation | 242 | 0.65 | 0.10 | 2 studies:<br>Proebstle et al, <sup>54</sup> 2008;<br>Kendler et al, <sup>55</sup> 2013 |
|  | Phlebectomy | 324 | 1.73 | 0.52 | 3 studies:<br>Krasznai et al, <sup>56</sup> 2015;<br>Smith et al, <sup>57</sup> 1998;<br>Creton et al, <sup>58</sup> 2012 |
| **Plastic**<br>**Surgery** | Liposuction/<br>Related<br>Procedures | 206 | 0.53 | 0.44 | 3 studies:<br>Schmeller et al, <sup>59</sup> 2012;<br>Augustin et al, <sup>60</sup> 2010;<br>Danilla et al, 2013 |
|  | Breast<br>Procedures | 20 | 2.6 | n/a | 2 studies:<br>Khater et al, <sup>22</sup> 2017 |
|  | Facial Surgery/<br>Cleft Lip<br>Surgery | 112 | 3.99 | 0.49 | 1 study:<br>Bertelsen et al, <sup>62</sup> 2011<br>Kim et al, 2017 |
| **Dermatology** | Ablative laser/<br>Electrochemoth<br>erapy | 12 | 1.27 | 0.20 | 2 studies:<br>Kessels et al, 2012<br>Kendler et al 2013 |
| **Other**<br>**Surgery** | Hand Surgery | 52 | 2.77 | 0.43 | 2 studies:<br>Prasetyono et al, <sup>63</sup> 2016;<br>Ceran et al, <sup>64</sup> 2015 |

### Complications reported in TLA cases (Table 3)

Table 3 summarizes the post-operative complications based on 38,430 cases using TLA. Liposuction was by far the most commonly reported procedure with 27,767 cases. Endovenous laser ablation and mastectomy were also well-reported at 3,026 and 2,602 cases, respectively. The large majority of reported procedures including liposuction and endovenous laser ablation had complication risks below 5%. Phlebotomy, non-liposuction body reconstruction/ recontouring, breast reduction, hand surgery and inguinal hernia repair had post-operative complication rates in the 5-10% range (2.2, 7.9, 6.2, 9.5, and 5.4%, respectively). Only mastectomy, hyperhidrosis and facial surgery had higher complication risks (20.8, 11.4, and 10.9%, respectively). In terms of specific complications experienced, only two occurred in more than 5% of cases: hematoma which occurred in 7.6% of 430 hyperhidrosis cases, and 5.4% of 612 inguinal hernia cases, and skin or fat necrosis which occurred in 13.2% of the 2,602 mastectomy cases reported.

**Table 3.**
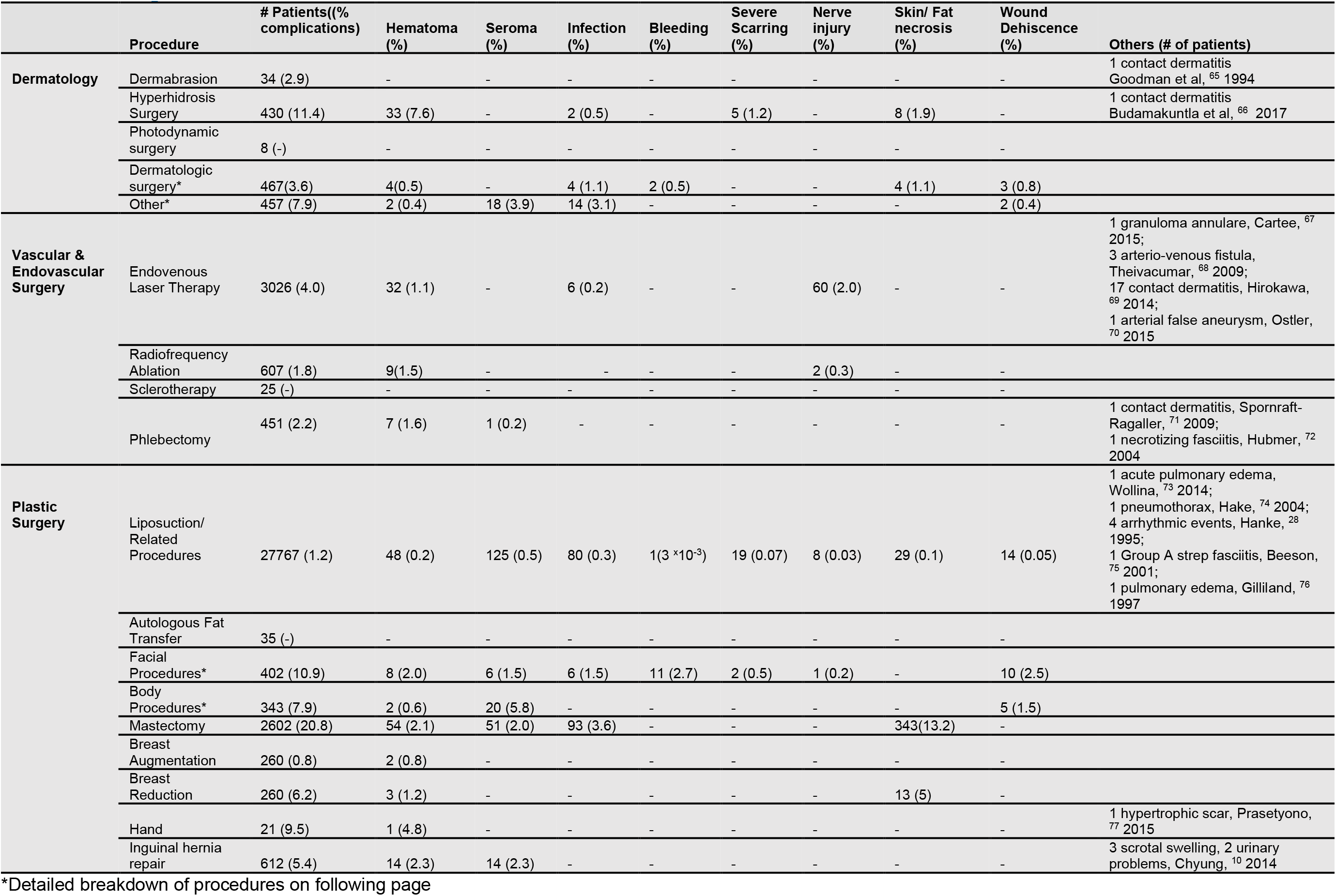
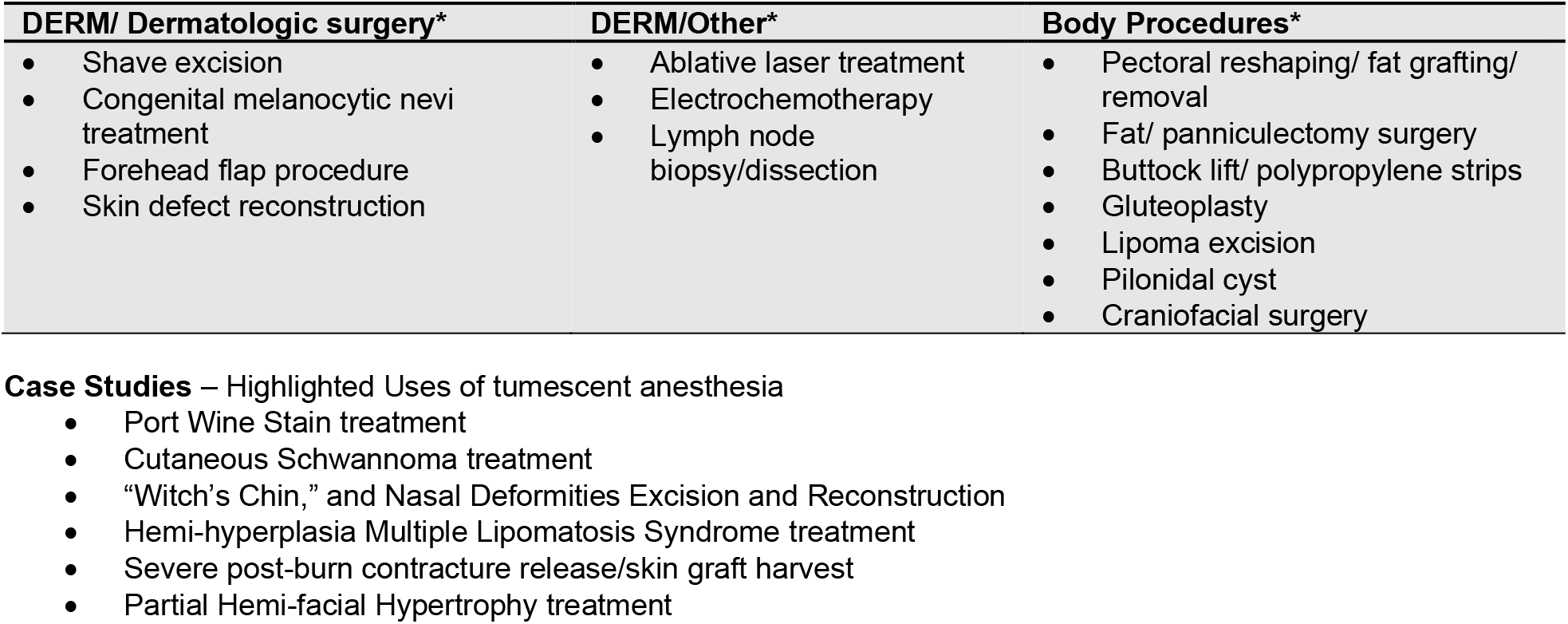
Complications of Tumescent Local Anesthesia.

|  | Procedure | # Patients(% complications) | Hematoma (%) | Seroma (%) | Infection (%) | Bleeding (%) | Severe Scarring (%) | Nerve injury (%) | Skin/ Fat necrosis (%) | Wound Dehiscence (%) | Others (# of patients) |
| --- | --- | --- | --- | --- | --- | --- | --- | --- | --- | --- | --- |
| **Dermatology** | Dermabrasion | 34 (2.9) | - | - | - | - | - | - | - | - | 1 contact dermatitis Goodman et al, <sup>65</sup> 1994 |
|  | Hyperhidrosis Surgery | 430 (11.4) | 33 (7.6) | - | 2 (0.5) | - | 5 (1.2) | - | 8 (1.9) | - | 1 contact dermatitis Budamakuntla et al, <sup>66</sup> 2017 |
|  | Photodynamic surgery | 8 (-) | - | - | - | - | - | - | - | - |  |
|  | Dermatologic surgery* | 467(3.6) | 4(0.5) | - | 4 (1.1) | 2 (0.5) | - | - | 4 (1.1) | 3 (0.8) |  |
|  | Other* | 457 (7.9) | 2 (0.4) | 18 (3.9) | 14 (3.1) | - | - | - | - | 2 (0.4) |  |
| **Vascular & Endovascular Surgery** | Endovenous Laser Therapy | 3026 (4.0) | 32 (1.1) | - | 6 (0.2) | - | - | 60 (2.0) | - | - | 1 granuloma annulare, Cartee, <sup>67</sup> 2015;<br>3 arterio-venous fistula, Theivacumar, <sup>68</sup> 2009;<br>17 contact dermatitis, Hirokawa, <sup>69</sup> 2014;<br>1 arterial false aneurysm, Ostler, <sup>70</sup> 2015 |
|  | Radiofrequency Ablation | 607 (1.8) | 9(1.5) | - | - | - | - | 2 (0.3) | - | - |  |
|  | Sclerotherapy | 25 (-) | - | - | - | - | - | - | - | - |  |
|  | Phlebectomy | 451 (2.2) | 7 (1.6) | 1 (0.2) | - | - | - | - | - | - | 1 contact dermatitis, Spornraft-Ragaller, <sup>71</sup> 2009;<br>1 necrotizing fasciitis, Hubmer, <sup>72</sup> 2004 |
| **Plastic Surgery** | Liposuction/ Related Procedures | 27767 (1.2) | 48 (0.2) | 125 (0.5) | 80 (0.3) | 1(3 x10<sup>-3</sup>) | 19 (0.07) | 8 (0.03) | 29 (0.1) | 14 (0.05) | 1 acute pulmonary edema, Wollina, <sup>73</sup> 2014;<br>1 pneumothorax, Hake, <sup>74</sup> 2004;<br>4 arrhythmic events, Hanke, <sup>28</sup> 1995;<br>1 Group A strep fasciitis, Beeson, <sup>75</sup> 2001;<br>1 pulmonary edema, Gilliland, <sup>76</sup> 1997 |
|  | Autologous Fat Transfer | 35 (-) | - | - | - | - | - | - | - | - |  |
|  | Facial Procedures* | 402 (10.9) | 8 (2.0) | 6 (1.5) | 6 (1.5) | 11 (2.7) | 2 (0.5) | 1 (0.2) | - | 10 (2.5) |  |
|  | Body Procedures* | 343 (7.9) | 2 (0.6) | 20 (5.8) | - | - | - | - | - | 5 (1.5) |  |
|  | Mastectomy | 2602 (20.8) | 54 (2.1) | 51 (2.0) | 93 (3.6) | - | - | - | 343(13.2) | - |  |
|  | Breast Augmentation | 260 (0.8) | 2 (0.8) | - | - | - | - | - | - | - |  |
|  | Breast Reduction | 260 (6.2) | 3 (1.2) | - | - | - | - | - | 13 (5) | - |  |
|  | Hand | 21 (9.5) | 1 (4.8) | - | - | - | - | - | - | - | 1 hypertrophic scar, Prasetyono, <sup>77</sup> 2015 |
|  | Inguinal hernia repair | 612 (5.4) | 14 (2.3) | 14 (2.3) | - | - | - | - | - | - | 3 scrotal swelling, 2 urinary problems, Chyung, <sup>10</sup> 2014 |

\*Detailed breakdown of procedures on following page

| DERM/ Dermatologic surgery* | DERM/Other* | Body Procedures* |
| --- | --- | --- |
| • Shave excision<br>• Congenital melanocytic nevi treatment<br>• Forehead flap procedure<br>• Skin defect reconstruction | • Ablative laser treatment<br>• Electrochemotherapy<br>• Lymph node biopsy/dissection | • Pectoral reshaping/ fat grafting/ removal<br>• Fat/ panniculectomy surgery<br>• Buttock lift/ polypropylene strips<br>• Gluteoplasty<br>• Lipoma excision<br>• Pilonidal cyst<br>• Craniofacial surgery |

**Case Studies** – Highlighted Uses of tumescent anesthesia
- Port Wine Stain treatment - Cutaneous Schwannoma treatment - “Witch’s Chin,” and Nasal Deformities Excision and Reconstruction - Hemi-hyperplasia Multiple Lipomatosis Syndrome treatment - Severe post-burn contracture release/skin graft harvest - Partial Hemi-facial Hypertrophy treatment

### Reported Cases of Death Associated with TLA Use

Thirteen cases of death have been reported in TLA procedures, all associated with liposuction performed before 2003. Rao and colleagues ^25^ reported 5 deaths associated with 48,527 tumescent liposuction procedures conducted between 1993-1998. Kaminer and colleague reported 4 deaths in tumescent liposuction cases conducted over 18 years from 1983-2001, but did not report total number of procedures performed.^26^ Lehnhardt et al reported 4 TLA-related deaths among 72 severe complications reported in a survery study (2,275 respondents) regarding liposuction procedures in Germany between 1998-2002.^27^ The causes of death in these 13 cases were TLA-related in 5 cases (lidocaine/bupivacaine toxicity in 4 cases, and fluid overload in a single case) and likely TLA-unrelated in 8 cases [pulmonary embolus (4 cases), concurrent general anesthetic complications (2 cases), hemorrhage (1 case), visceral perforation (1 case)]. There have been no deaths in the 33,429 TLA cases published in 146 studies since 2003.^30-32^ These above studies collectively indicate that TLA is a safe procedure.

## DISCUSSION

The findings herein reflect a body of evidence that TLA has an excellent safety profile and effectively controls intraoperative and post-operative pain, with less complications. Mean pain scores ranged from 0.53-3.99 on a 0-10 analog scale indicating minimal pain experienced intra-operatively and on post-operative day 1 (see Table 1). In addition, TLA may be safer than general anesthesia. When liposuction was introduced into clinical practice by Giorgio Fischer in the 1970s,^31,33^ serious complications were common and the greatest risks were associated with general anesthesia.^31^ Application of the TLA was an important technical breakthrough as it can avoid the most severe complications encountered in the early years of liposuction with general anesthesia.^1,2^ Though five TLA-related deaths have been reported, all were in liposuction cases performed prior to 2003 when maximal lidocaine doses and fluid volumes were still being established. There have been no deaths in the 33,429 TLA cases published since 2003.

In the past three decades, TLA anesthesia has become a standard anesthetic approach with good pain management and low complication risks.^1-5,24^ The technical advancement and standardization of TLA has paved the way for expansion of its application beyond liposuction to various other minimally invasive surgical procedures involving cutaneous and subcutaneous tissues (e.g. breast, vascular, hand, hernia, and cleft-palate surgery).^4,5,24^ In these procedures, TLA has shown advantages not only in efficacy and safety, but also in terms of cost reduction in its application for office-based surgical procedures.

The results herein report a subset of procedures in which TLA had a relatively higher complication risk, i.e. mastectomy (20.8%), hyperhidrosis surgery (11.4%) and facial surgery (10.9%). Comparable data regarding complications in similar surgeries performed under general anesthesia is only available for breast surgery. Risk of flap necrosis in breast reconstruction has been reported to be 14% with general anesthesia which is comparable to the 12.8% risk reported herein with TLA in a retrospective study of total 730 cases with breast reconstructions.^11^ Similarly, risk of skin necrosis in mastectomy appears to be comparable in general anesthesia vs. TMA cases (7% vs 10%, respectively) in a retrospective study which included 601 nontumescent and 890 tumescent cases.^9^ However, two smaller studies containing 100 and 332 tumescent patients, respectively, reported the risk of skin necrosis in mastectomy to be 34% and 15% higher with tumescent anesthesia than general anesthesia, respectively.^34,35^ Using lower dose of lidocaine (not exceeding 28 mg/kg) in the TLA might be helpful. In their daily clinical practice, many surgeons may, however, associate TLA with deep sedation or general anesthesia in some procedures, i.e. abdominoplasty, mastectomy, or large volume liposuction. In addition to the above mentioned risk or side effects, the total volume of the infiltrated liquid during the TLA procedure should be carefully regulated, as the fluid overload may result in heart failure.

Long-acting local anesthetics such as liposomal bupivicaine injected locally at the end of joint replacement surgery have been recently associated with outstanding pain control and no elevation in complications. These formulations contain much higher concentrations of bupivicaine and epinephrine than TLA but appear to be largely devoid of complications.^36^ Additional approaches for optimization of pain management during the TLA procedures include 1) slowing rate of infiltration, 2) vibrating the skin, 3) warming TLA solution, and 4) cooling the local skin of the TLA procedure,^4,37^ as well as 5) using thinner needles, and 6) adjusting needle insertion angles.^38^ Zelickson and colleagues reported the parallel and minimal needle-insertion technique to the skin into the superficial dermis with 0.5% lidocaine containing 1:200,000 epinephrine buffered 1:10 with 8.4% sodium bicarbonate achieved a painless injection of local anesthetic.^38^

The cost for office-based surgery with TLA is much lower than the same surgical procedures conducted as inpatients with general anesthesia.^39^ Broader application of TLA would therefore reduce costs and possibly reduce hospital-based iatrogenic morbidity, i.e. hospital acquired infection. However, given the inherent limitation of TLA application to cutaneous and subcutaneous tissues where local infiltration of anesthetics can be easily achieved, TLA is not practical for intrathoracic, abdominal, or neural surgeries, or for prolonged or highly complex superficial surgeries.

## CONCLUSIONS

The data herein indicate that TLA has an outstanding record of safety and efficacy across a wide range of surgical procedures. TLA safety appears to be equivalent or superior to general anesthesia with minimal patient discomfort/complications and very low risk of death. Its low-cost and rapid patient recovery warrant further studies of cost-reduction and patient satisfaction. Expanded education of TLA techniques in surgical and anesthesia training programs should be undertaken in order to broaden patient access to this safe and effective anesthetic modality for cutaneous and subcutaneous surgical procedures.

## Data Availability

This is a systematic review article based on previously published works and data spanning the past 30 years (1990 - 2019). This work includes data analysis from 157 original clinical research articles.

